# A cross-sectional cohort study of prevalence of antibodies to COVID-19 in Port-au-Prince, Haiti

**DOI:** 10.1101/2021.06.30.21259815

**Authors:** Vincent DeGennaro, Timothy Schwartz, Rebecca Henderson, Marie-Carmelle Elie

**Affiliations:** Innovating Health International, Port au Prince, Haiti; Socio-Dig Research, Port-au-Prince, Haiti; Department of Medicine, University of Florida College of Medicine, Gainesville, FL

**Keywords:** COVID-19, Haiti, Infectious Disease, Virus, low-income country

## Abstract

**Intro:** As of January 14, 2021, Haiti has had 10,781 confirmed (first case March 19th) and 45,927 suspected cases of COVID-19, with 240 official deaths.

**Methods:** From May until September, 2020, we tested visitors to 20 clinics, for COVID-19 in five neighborhoods of Port-au-Prince as part of a public health effort to determine prevalence of COVID-19 in the general community. In order to estimate changes in number of deaths, the team visited eighteen funeral homes to solicit data on the number of funerals conducted for each month in 2019 and through October 2020. We also sought to evaluate the attitudes of Port-au-Prince citizens towards a generic COVID-19 vaccine in April 2021.

**Results:** In May and July 2020, 11.4% and 9.1% of those tested were positive for antibodies to COVID-19, respectively. The number of funerals held in the Port-au-Prince area increased by 69.6.% (CI 95% 56.1-83.1) since the official arrival of COVID-19 on March 19^th^. We found high rates of vaccine hesitancy with 76% saying they would not take a free COVID-19 vaccine. Further research is needed to validate the findings here, but there are strong suggestions that COVID-19 has had more of an impact than previously reported.

**‘What is already known on this subject?’:** Very little is known about the true epidemiology of COVID-19 in Haiti due to lack of testing, stigma, and lack of resources. The Ministry of Health reports Haiti has had 10,781 confirmed (first case March 19th) and 45,927 suspected cases of COVID-19, with 240 official deaths. Most officials accept these numbers as vast underestimates.

**‘What does this study add?’’:** This study estimates a prevalence of 10% for COVID-19 antibodies in the population of Port-au-Prince in May to August 2020. Only 19-38% of those surveyed with confirmed antibodies reported experiencing symptoms in the last few months prior to the survey. We found a 69% increase in funerals per month in Port-au-Prince for the first six months post-COVID (March 2020) compared to the 14 months prior.

Finally, we found significant vaccine hesitancy with only 24% reporting “Yes” or “Maybe” when asked if they’d take a free of charge vaccine.

## Intro

In Haiti, as in much of the developing world, official numbers of confirmed COVID-19 and mortality from the disease remains relatively low. As of January 14, 2021, Haiti has had 10,781 confirmed (first case March 19^th^) and 45,927 suspected cases of COVID-19, with 240 deaths (1). However, tracking COVID-19 in developing nations poses a logistical challenge given limited testing capacity, lack of access, and poor health literacy. We present our findings following the implementation of serologic testing performed at two timepoints in Haiti during the epidemic. We also present public data on deaths in funeral rates collected from funeral homes and attitudes towards a COVID-19 vaccine in Haiti.

## Methods

From May 20 until September 1, 2020, we tested visitors to 20 clinics, including staff and family members of patients, for COVID-19 in five neighborhoods of Port-au-Prince, Haiti as part of a public health effort to determine prevalence of COVID-19 in the general community. All visitors were over 18 years old, including patients and family, were approached prior to entering the clinics for consent to a brief survey and a COVID-19 antibody test through a fingerstick.

Participants provided demographic and symptomatic information as part of a structured checklist verbally in Haitian Kreyol to study personnel. After completing the brief questionnaire, participants underwent fingerpick blood testing using the API Pharma Covid-Rapid point of care serology test kit (2), according to company instructions. Results were communicated to participants, and clinical guidance, including the need to self-isolate and to seek medical care if symptoms worsen, were provided for those testing positive for active infection as determined by the presence of IgM antibodies.

After the first 289 participants were tested over three weeks in May, an overall prevalence of infection was ascertained. Testing was then paused for three weeks to allow detection of change in prevalence over time. Testing resumed in July, and the procedures were repeated for the first 288 clinic visitors. We present point prevalence data for six neighborhoods of Port-au-Prince at two distinct time points at the height of the first wave of the COVID-19 epidemic.

In order to estimate changes in number of deaths, the team visited eighteen funeral homes to solicit data on the number of funerals conducted for each month in 2019 and through October 2020. All eighteen funeral homes agreed to participate and they are from five distinct geographical regions of the metropolitan area, including the furthest North, South, and West as well as the heart of the city. No other demographic data on the funerals themselves was collected.

We also sought to evaluate the attitudes of Port-au-Prince citizens towards a generic COVID-19 vaccine in April 2021, posing the question based on the assumption that it was provided free of charge. We attempted to contact by phone 140 people selected at random from the original May 2020 cohort. We succeeded in getting in touch with 88 (63%), which is a similar rate of response to other studies conducted by phone in Haiti by Sociodig when not using a recontact networking strategy. Six of the 52 non-respondents said that a new person now had the phone number and the rest rang with no response.

## Results

In May and July 2020, 11.4% and 9.1% of those tested were positive for antibodies to COVID-19 (Table 1), respectively. The incidence of COVID-like symptoms among participants or household contacts did not change significantly with regards to their antibody status, with the range of those reporting COVID-like symptoms in themselves or household contacts being from 19-38%. Rates of antibody presence were relatively similar regardless of age, sex, education level, or neighborhood.

**Table 1.** Antibody Test Results and Symptoms at Two Timepoints

|  | May 2020 | July 2020 | Overall |  |
| --- | --- | --- | --- | --- |
|  | Results | Results | Participant with recent symptoms | Participant with household contacts with recent symptoms |
| **IgM and IgG** | 1% (n=3) | 1.7% (n=5) | 38% (n=8) | 20% (n=5) |
| **IgG only** | 10.3% (n=30) | 7.3% (n=21) | 22% (n=51) | 30% (n=10) |
| **All positives** | 11.4% (n=33) | 9.0% (n=26) | 23% (n=59) | 27% (n=15) |
| **Negative** | 88.6% (n=256) | 90.9% (n=215) |  |  |
| **All participants: May** |  |  | 78 (27%) | 110 (38%) |
| **All participants: July** |  |  | 46 (19%) | 54 (22%) |
| **Total** | N=289 | N= 241 | N=530 | N=530 |

We compared average monthly rates of funerals reported by 18 funeral homes between 2019 and 2020 (Table 2). we found a statistically significant increase in the rate of funerals at each month starting in March 2020. The number of funerals held in the Port-au-Prince area has increased by 69.6.% (CI 95% 56.1-83.1) since the official arrival of COVID-19 on March 19^th^.

**Table 2.** Monthly comparison of mean number of funerals across 18 funeral homes, 2019-2020

|  | Jan | Feb | March | April | May | June | July | Aug | Sept | Oct | Nov | Dec |
| --- | --- | --- | --- | --- | --- | --- | --- | --- | --- | --- | --- | --- |
| 2019 | 7.61 | 8.50 | 10.11 | 10.78 | 12.61 | 11.83 | 12.22 | 12.61 | 12.78 | 12.83 | 13.56 | 18.39 |
| 2020 | 13.17 | 10.83 | 13.67 | 17.11 | 19.22 | 21.00 | 18.89 | 17.78 | 15.28 | 18.00 |  |  |
|  |  | Mean Monthly<br>Funerals/<br>Funeral Home | Change |  | T-value |  | P value |  |  |  |  |  |
| Pre-COVID<br>funerals |  | 11.99<br>[CI 95% 10.707–<br>13.273] | + 69.6% (CI<br>95% 56.1-83.1) |  | 5.51 |  | .000011 |  |  |  |  |  |
| Post-COVID<br>funerals |  | 17.62<br>[CI 95% 16.026–<br>19.214] |  |  |  |  |  |  |  |  |  |  |

Six of the 51 said it was a wrong number and the rest rang but no response. After speaking on the phone with 88 of the participants from the May 2020 cohort, there were high rates of vaccine hesitancy. Of the 88 participants, only six (7%) said “Yes”, while 15 (17%) said “Maybe” and 67 (76%) said “No”.

## Discussion

Our results suggest that the prevalence and mortality of COVID-19 in Port-au-Prince have been underestimated due to under resourced public health surveillance infrastructure. We offer two separate mortality estimates based on our data. First, with an estimated population of 2.8 million people (3) and an approximately 10% prevalence by July, this means that 280,000 Port-au-Prince residents may have had COVID-19. Assuming a low-end range of 0.02-0.05% mortality rate (4) based on Haiti’s young age stratification, that is 5600-14,000 deaths. Second, Port-au-Prince reports approximately 23,282 deaths per year (5). Based on the 69.6% (CI 95% 56.1-83.1) increase in funerals from our survey of funeral homes, we estimate 16,204 (CI 95% 13,061-19,347) excess deaths possibly due to COVID-19 in Port-au-Prince alone. Other factors could have contributed to the change in funeral home usage, but we consider this unlikely given the correspondence between COVID-19’s presence in Haiti and the increase in funerals in Port-au-Prince. Many smaller funeral homes did not have variability in rates over time; future work would recommend assessing those areas specifically to determine if there was regional variation.

The official MSPP statistics say that there have been 240 deaths in Port-au-Prince due to COVID-19 as of January 14^th^ 2020, with a 2.23% mortality rate (1), at least one and quite possibly two orders of magnitude higher than noted elsewhere in the world. This could be due to poor healthcare infrastructure or more likely, significant undertesting meaning the confirmed cases are much more likely to overrepresent the very sick. There have been persistent problems with widespread COVID-19 testing; including that the supply chain for testing supplies is frequently disrupted and rumors circulated that testing could lead to becoming infected or being taken from your family if you test positive.

We used API Pharma’s Covid-Rapid Antibody test, which has a 94% sensitivity and 100% specificity overall. For those in the first 14 days after infection, sensitivity can be as low as 61-83%. As our cohort was largely asymptomatic and did not report exposure, it is impossible to know if we are missing early cases of COVID-19. More importantly, the 100% specificity means that this sample likely represents an underestimate of prevalence since all positives are true positives but not all negatives are true negatives.

Trust in modern medicine has variable penetration across Haiti, with the highest rates in Port-au-Prince. While the cohort contacted to discuss their willingness to take the vaccine was small, it is nonetheless of significant concern that three out of four participants would decline a free COVID-19 vaccine at present in Port-au-Prince, the city that likely saw the most COVID-19 morbidity and mortality and also has the highest levels of education. Further research would be needed to evaluate if the low official mortality figures play a role in willingness to take a vaccine and if the high numbers of deaths from this study might convince more people to take the vaccine.

## Data Availability

Data is available upon request.

## Acknowledgements

Many thanks to the staff at Innovating Health International and Sociodig for performing this research.

